# A multi-center retrospective chart analysis identifying clinical, biochemical, and radiographic signatures of short-chain enoyl-CoA hydratase (ECHS1) deficiency

**DOI:** 10.64898/2026.11.05.26366953

**Authors:** Christine Kong, Cesar Alves, Georg-Friedrich Vogel, Rui Xiao, Irina Anselm, Gerard Berry, Juan Francisco Cabello, Verónica Cornejo, Andrea Cortes Fernandez, Gregory M. Enns, Michael Gomez, Chad Haldeman-Englert, Suzanne Hollander, A. Micheil Innes, Emma Jannotta, Aaina Kochhar, Jessica Kopesky, Austin Larson, Chung Lee, María Lezcano, Rachel Li, Matthew Lines, Jennifer Morrison, Michael Muriello, Elizabeth Null, Luis Peña-Quintana, Erin Phelps, Mohamad Saifeddine, Marta Cristina Sanabria, Russell P. Saneto, Emily Shelkowitz, Christina G. Tise, Johan L.K. Van Hove, Ibrahim Elsharkawi, Rebecca Ganetzky

**Affiliations:** Perelman School of Medicine, University of Pennsylvania, Philadelphia, Pennsylvania, USA; Mitochondrial Medicine Frontier Program, Division of Human Genetics, Department of Pediatrics, The Children’s Hospital of Philadelphia, Philadelphia, Pennsylvania, USA; Division of Neuroradiology, Department of Radiology, The Children’s Hospital of Philadelphia, Philadelphia, Pennsylvania, USA; Division of Neuroradiology, Department of Radiology, Boston Children’s Hospital and Harvard Medical School, Boston, Massachusetts, USA; Department of Paediatrics I and Institute of Cell Biology, Medical University of Innsbruck, Innsbruck, Austria; X Institute of Cell Biology, Medical University of Innsbruck, Innsbruck, Austria; Department of Neurology, Boston Children’s Hospital and Harvard University, Boston, Massachusetts, USA; Division of Genetics and Genomics, Department of Pediatrics, Boston Children’s Hospital and Harvard University, Boston, Massachusetts, USA; Instituto de Nutricion y Tecnologia de los Alimentos (INTA), Universidad de Chile, Santiago, Chile; Division of Human Genetics, Department of Pediatrics, Cincinnati Children’s Hospital Medical Center and University of Cincinnati, College of Medicine, Cincinnati, OH, USA; Division of Medical Genetics, Department of Pediatrics, Lucile Packard Children’s Hospital and Stanford University, Stanford, California, USA; Pediatrix Medical Group of Florida at Orlando Health, Winnie Palmer Hospital for Women and Babies, Orlando Health, Orlando, Florida, USA; Cone Health Precision Health, Greensboro, North Carolina, USA; Department of Medical Genetics, Cumming School of Medicine, University of Calgary, Calgary, Alberta, Canada; Section of Clinical Genetics and Metabolism, Department of Pediatrics, Children’s Hospital Colorado and University of Colorado, Aurora, Colorado, USA; Clinical Nutrition Department, Children’s Wisconsin, Milwaukee, Wisconsin, USA; Facultad de Ciencias Médicas, Universidad Nacional de Asunción, San Lorenzo, Paraguay; Department of Pediatrics, Division of Medical Genetics, Sanford Health and Sanford School of Medicine, University of South Dakota, Sioux Falls, South Dakota, USA; Division of Genetics, Department of Pediatrics, Children’s Nebraska and University of Nebraska Medical Center, Omaha, Nebraska, USA; Pediatric Gastroenterology and Nutrition Unit, Insular-Materno Infantil University Hospital Complex, CIBEROBN-ISCIII, University of Las Palmas de Gran Canaria, Las Palmas de Gran Canaria, Spain; Clinical Nutrition Department, Seattle Children’s Hospital, Seattle, Washington, USA; Department of Pediatrics, Division of Pediatric Neurology, Sanford School of Medicine, University of South Dakota, Sioux Falls, South Dakota, USA; Neuroscience Institute, Norcliffe Center for Integrated Brain Research, Division of Pediatric Neurology, Department of Neurology, Seattle Children’s Hospital and University of Washington, Seattle, Washington, USA; Division of Genetic Medicine, Department of Pediatrics, Seattle Children’s Hospital and University of Washington, Seattle, Washington, USA; Division of Medical Genetics, Department of Genetics and Genomic Sciences, Icahn School of Medicine, New York, New York, USA

**Keywords:** ECHS1 deficiency, mitochondrial disease, inborn errors of metabolism, genotype-phenotype, valine metabolism

## Abstract

Short-chain enoyl-CoA hydratase (ECHS1) catalyzes the hydration of C4-C6 acyl-CoA molecules to produce ATP in fatty acid β-oxidation. Deficiency of the ECHS1 enzyme, ECHS1D, is a rare autosomal recessive disorder that presents with a large and pleomorphic clinical spectrum. We present the largest comprehensive study of known diagnosed ECHS1D patients to date, reporting 26 new patients and 8 novel variants in conjunction with a literature review of 87 previously reported patients. All patients were categorized into neonatal, infantile, and late-onset groups based on age of onset and clinical course. Neonatal-onset cases were characterized by early and severe multisystem disease, often with lactic acidosis, hypotonia, cardiomyopathy, and high early mortality (62.5%). The infantile-onset group was heterogeneous, but many exhibited hallmark features such as dystonia, developmental delay, and basal ganglia involvement on neuroimaging. Mortality was significantly lower than the neonatal cohort (24.4%). Late-onset patients had more localized neurologic symptoms, particularly dystonia, and significantly lower mortality overall (0%). Moreover, we report a 37.5% survival rate in newly reported neonatal onset patients compared with 18.2% in prior reports, possibly reflective of earlier recognition of ECHS1D and greater utilization of therapies including N-acetylcysteine, sodium dichloroacetate, and valine-restricted diet.

## INTRODUCTION

Mitochondria are the primary cellular source of ATP production through oxidative phosphorylation (OXPHOS) and fatty acid β-oxidation. In the mitochondria, short-chain enoyl-CoA hydratase (ECHS1, or SECH) was originally identified to catalyze the second step of fatty acid β-oxidation, the hydration of C4-C6 fatty acyl-CoA molecules, to produce ATP. More recently, it was discovered that perhaps more importantly than its role in fatty acid β-oxidation, ECHS1 has a critical function in valine catabolism, metabolizing and detoxifying highly reactive byproducts of valine oxidation, methacrylyl-CoA and acryloyl-CoA.^1,2^ It is this second role whose impairment seems to be central to causing monogenic disease. The ECHS1 enzyme is encoded by the *ECHS1* gene, found on chromosome 10q26.2-26.3 and localized in the mitochondrial matrix as a 160 kD homohexameric enzyme.^3,4^ Deficiency of the ECHS1 enzyme (ECHS1D) has been identified as a rare autosomal recessive inborn metabolic disorder.

Biochemically classifying ECHS1D is challenging. Clinically, ECHS1D presents like a primary mitochondrial disease rather than a disorder of fatty acid β-oxidation, with combined respiratory chain deficiency and secondary pyruvate dehydrogenase deficiency^4^, leading to Leigh Syndrome and a broad phenotypic spectrum.^5^ More specifically, across the large and pleomorphic clinical spectrum of ECHS1D phenotypes, there have been three previously reported broad phenotype groupings^1^:

1. Neonatal presentation with severe lactic acidosis, neonatal seizures, encephalopathy, hypotonia. Patients with this presentation typically have rapid disease progression and high mortality rates.
2. A Leigh syndrome presentation, typically in the infantile period with global developmental regression, resulting in severe developmental delay and death.
3. Late-onset presentation in infancy or childhood with possible developmental delay and an invariant feature of dystonia, typically a paroxysmal dystonia.

Biochemical and clinical parameters alone may be insufficiently specific and sensitive for diagnosing ECHS1D. Molecular testing is relied upon for definitive diagnosis, but even so, many individuals do not undergo molecular testing until much later in life or even posthumously.^6^ There have been efforts to explore the full clinical picture of ECHS1D,^7,8^ however, most cases are known via individual case presentations.

Thus, the objective of this study was to perform a comprehensive study of all the known diagnosed ECHS1D patients to identify the most likely clinical, biochemical, and radiographic markers for diagnosis and most common phenotypes, and assess possible benefit of various therapeutic strategies. We also sought to identify clearer markers of grouping for ECHS1D diagnoses to better elucidate the phenotypic spectrum of the disease.

Here, we report 26 new patients with ECHS1D, including 8 novel variants. We also conducted a literature review of the 87 previously reported patients with ECHS1D, bringing the total of known patients to 113, 33 of whom are deceased as known to date. To our knowledge, this is the largest known collection of ECHS1D patients and allows for the development of generalized information about this rare mitochondrial disease.

## METHODS

### Study Design and Data Acquisition

This is a multi-center, retrospective, cross-sectional study of individuals diagnosed with ECHS1D. Patient data were collected through collaboration with multiple international metabolic and genetic clinics, research institutions, and published case reports.

Previously reported patients with a confirmed molecular diagnosis of ECHS1D were identified through a systematic review of the literature using PubMed with the search terms “ECHS1”, “ECHS1 deficiency”, and “short-chain enoyl-CoA hydratase deficiency.” Inclusion criteria for published patients included documentation of biallelic pathogenic or likely pathogenic variants in the *ECHS1* gene and sufficient clinical, neuroimaging, or biochemical data to allow for phenotype classification.

New, unpublished cases were collected from contributing institutions. Collaboration was also solicited via e-mail through the international “Metab-L” listserv. Inclusion criteria for new patients included clinical suspicion of a mitochondrial disorder, followed by genetic confirmation of biallelic *ECHS1* variants.

For all patients (published and unpublished), we extracted demographic data (age, sex, ancestry), clinical features (e.g., dystonia, hypotonia, seizures, lactic acidosis), biochemical findings (e.g., elevated lactate, valine metabolites, respiratory chain enzyme activities), neuroimaging findings (MRI), molecular genetic results, and treatment interventions (e.g., valine-restricted diets, N-acetylcysteine up until their death or last date of follow-up before 6/1/2024.)

Clinical presentations were grouped into three broad phenotypes (neonatal, infantile, and childhood-onset) based on age of onset. While age of onset is not a perfect metric for categorizing patients with ECHS1D, based on the large number of patients in the study, we deemed it most suitable for a broad categorization in a disease in which long-term functional and survival outcomes are not perfectly understood.

Neuroimaging review was performed by a central pediatric neuroradiologist and included an analysis of previously published data alongside a detailed assessment of new, unpublished cases involving patients with confirmed *ECHS1* causative variants. Where possible, MRI images themselves were reviewed (N=10); where not available, MRI reports were reviewed (N=83). Patterns were grouped based on the presence of malformative features, such as corpus callosum malformation, ventricular system abnormalities (including dilatation and septation of the lateral ventricles), and parenchymal volume loss, as well as features suggestive of chronic or acute/active lesions. These were characterized by volume loss or an edematous appearance, with or without restricted diffusion on diffusion-weighted imaging (DWI) when available, involving the cortex, white matter, basal ganglia, brainstem, and cerebellum.

### Population Frequency Estimate

gnomAD v4.1.1. population frequency data was downloaded for all *ECHS1* variants with predicted loss of function by gnomAD.^9^

### Variant Interrogation

Variant pathogenicity was further assessed using *in silico* tools and cross-referenced with gnomAD population frequency data.^9^ SIFT and PolyPhen-2 scores were calculated for all newly reported missense variants.^10,11^ For newly reported variants available on gnomAD, CADD scores were derived from the variant-specific gnomAD page.

### Statistics and Software

Density plots were generated using the *ggplot2* package, and world maps were created using the *ggplot2* and *maps* packages in R Statistical Software (v4.3.1; R Core Team 2023). Schematics and figures were compiled using Illustrator CC 2019 (Adobe).

Categorical outcomes (death, clinical category, lactic acidosis, seizures/epilepsy, and cardiomyopathy) were compared between variant categories using Fisher’s exact test. Association between age at presentation and age at death with mutation category were assessed using linear regression models. Mutation categories with insufficient sample size were excluded from analyses. The association of age of onset with age of death and age at presentation was assessed using Kaplan-Meier curves followed by log rank test. Age at last follow-up among surviving patients was compared between groups using Welch’s t-test.

### Ethics Review

This study was evaluated by the Children’s Hospital of Philadelphia Institutional Review Board and found to be an exempt retrospective protocol. Cases were recorded in a deidentified manner from all institutions. Ethical approval and data-sharing agreements were obtained from all participating centers in accordance with institutional guidelines.

## RESULTS

### Cohort Description

Our newly reported cohort includes 26 patients from 12 different institutions. Our systematic review identified 87 previously reported patients, for a total of 113 patients (Table 1). There are two newly reported sets of siblings for a total of 14 groups of siblings within our cohort (i.e. 99 unique pedigrees).^2, 12, 13, 14, 15, 16, 17, 18, 19, 20, 21, 22^

**Table 1.** Demographics.

|  | **NEWLY REPORTED COHORT**<br>(*N=26*) |  | **PREVIOUSLY REPORTED COHORT**<br>(*N=87*) |  |
|  | **Number** | **%** | **Number** | **%** |
|---|---|---|---|---|
| **MALE** | 14 | 53.8 | 45 | 51.7 |
| **FEMALE** | 12 | 46.2 | 42 | 48.3 |
| **ALIVE** | 20 | 77 | 59 | 69 |
| **DECEASED** | 6 | 23 | 27 | 31 |
| ***NEONATAL PRESENTATION*** *(N=30)* | 8 | 31 | 22 | 24.1 |
| **ALIVE** | 3 | 37.5 | 4 | 18.2 |
| **DECEASED** | 5 | 62.5 | 18 | 81.8 |
| **AVG. AGE OF PRESENTATION (MONTHS)** | 0.03 |  | 0.10 |  |
| **AVG. AGE OF DEATH (MONTHS)** | 5.1 |  | 10.5 |  |
| **MEDIAN AGE OF DEATH (MONTHS)** | 7 |  | 1.5 |  |
| **AVG. AGE AT LAST FOLLOW-UP FOR ALIVE PATIENTS (MONTHS)** | 52.67 |  | 81.9 |  |
| **MEDIAN AGE AT LAST FOLLOW-UP FOR ALIVE PATIENTS (MONTHS)** | 48 |  | 78 |  |
| ***INFANTILE PRESENTATION*** *(N=45)* | 8 | 31 | 37 | 42.5 |
| **ALIVE** | 7 | 87.5 | 28 | 75.7 |
| **DECEASED** | 1 | 12.5 | 9 | 24.3 |
| **AVG. AGE OF PRESENTATION (MONTHS)** | 7.4 |  | 6.55 |  |
| **AVG. AGE OF DEATH (MONTHS)** | 14 |  | 54.22 |  |
| **MEDIAN AGE OF DEATH (MONTHS)** | 14 |  | 28 |  |
| **AVG. AGE AT LAST FOLLOW-UP FOR ALIVE PATIENTS (MONTHS)** | 60.86 |  | 96.24 |  |
| **MEDIAN AGE AT LAST FOLLOW-UP FOR ALIVE PATIENTS (MONTHS)** | 36 |  | 84 |  |
| ***LATE ONSET*** *(N=38)* | 10 | 38 | 28 | 32.2 |
| **ALIVE** | 10 |  | 28 |  |
| **DECEASED** | 0 |  | 0 |  |
| **AVG. AGE OF PRESENTATION (MONTHS)** | 29.1 |  | 46.25* |  |
| **AVG. AGE AT LAST FOLLOW-UP FOR ALIVE PATEINTS (MONTHS)** | 135.6 |  | 162.34^ |  |
| **MEDIAN AGE AT LAST FOLLOW-UP FOR ALIVE PATIENTS (MONTHS)** | 108 |  | 135^ |  |

\*4/28 previously reported patients in the late onset category did not have their age of presentation reported
^10/28 previously reported patients in the late onset category did not have their age at last follow-up reported

### Population Genetics

We attempted to predict true population frequency of ECHS1D from population frequencies of pLOF variants in *ECHS1* per-gnomAD. GnomAD identified 62 pLOF variants, of which 4 had been classified pathogenic, 3 likely pathogenic, 1 a variant of uncertain significance and 2 with conflicting classifications.^9^ The most frequent allele with pLOF was p.Gly42Glufs*3, a reported pathogenic variant, with an allele frequency of 1.1*10^-4^. Collectively, assuming no person in GnomAD had multiple pathogenic alleles, the total allele frequency was 2.3*10^-4^. Assuming Hardy-Weinberg equilibrium, that predicts a disease frequency of 1.3*10^-8^, or approximately 1 per 77,000,000 people. As this would predict 4 incident cases in the United States we felt that this was likely an underestimate due to an overly conservative pLOF algorithm. Therefore, as an upper bound, we performed the same calculation from all pLOF or missense variants, of which there are 467. Of those, the ones that were classified as benign/likely benign (4) were removed. Of the remaining 463 variants, they had a collective allele frequency of 2.8*10 , predicting a disease frequency of 2.0*10 , or approximately 1/500,000. Of note, the common pathogenic c.489G>A variant, which has an allele frequency of 4.0*10 , was not included by GnomAD as a pLOF variant.

#### Genetics

1/26 newly reported patients had homozygous and 25/26 had compound heterozygous variants in the ECHS1 gene. 8/26 patients had at least one novel variant that has not previously been reported in the literature to our knowledge (Table 2). The full list of variants in newly reported patients is shared in Supplemental Table 1. Figure A depicts all ECHS1 variants reported in this study, indicated by black lines to the corresponding amino acid position. ECHS1 protein domains and regions of specialized function are highlighted. Figure B is a density plot with all ECHS1 variants reported in this study with respect to protein domains. We found that in our cohort, variants between amino acid position 155 and 195 are particularly frequent, portrayed by the inverted peak in figure B.

**Figure A.**
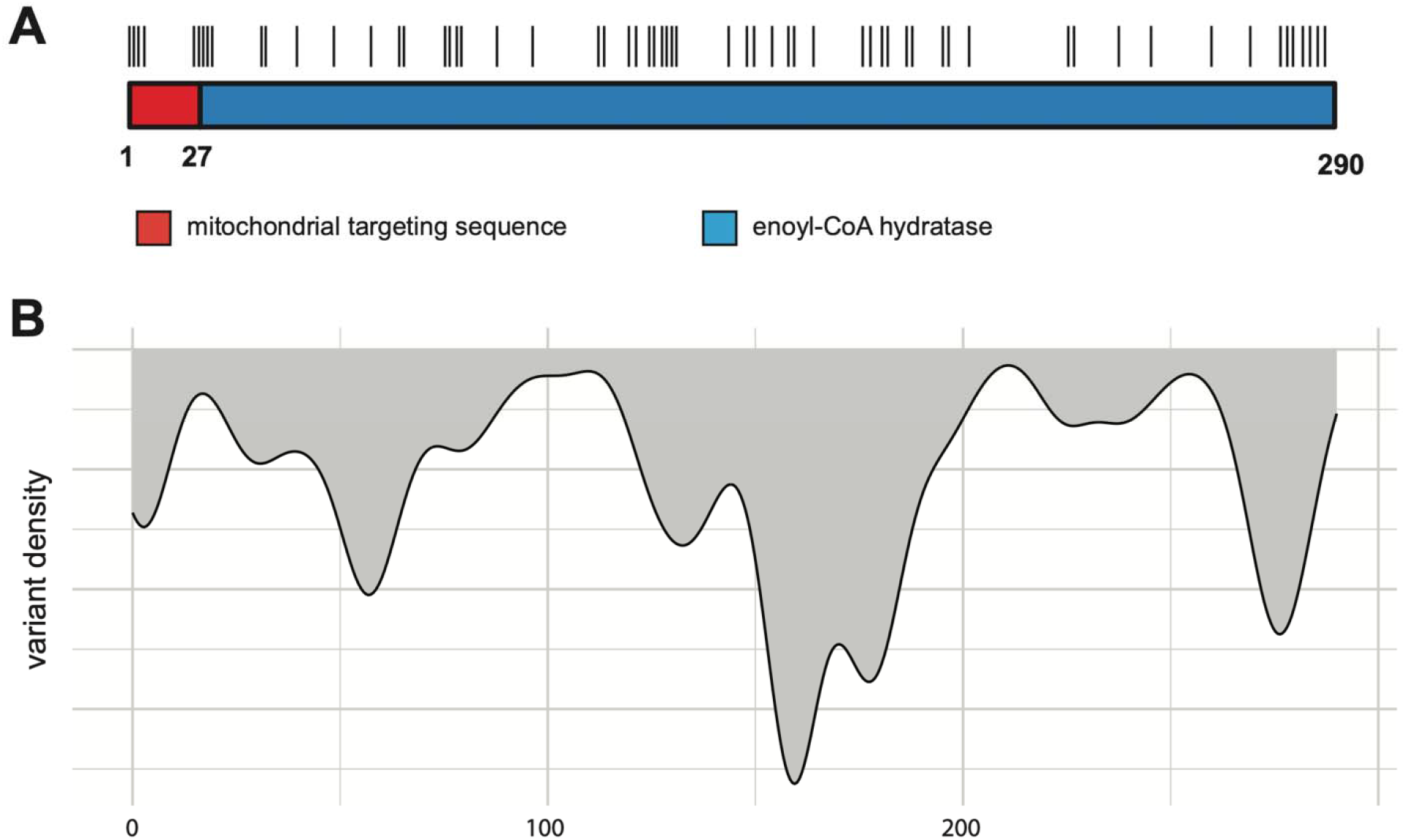
All *ECHS1* variants of this study are indicated by black lines to the corresponding amino acids position. *ECHS1* protein domains and regions of specialized function are highlighted. **Figure B.** Density plot *ECHS1* variants reported in this study with respect to protein domains.

**Table 2.**
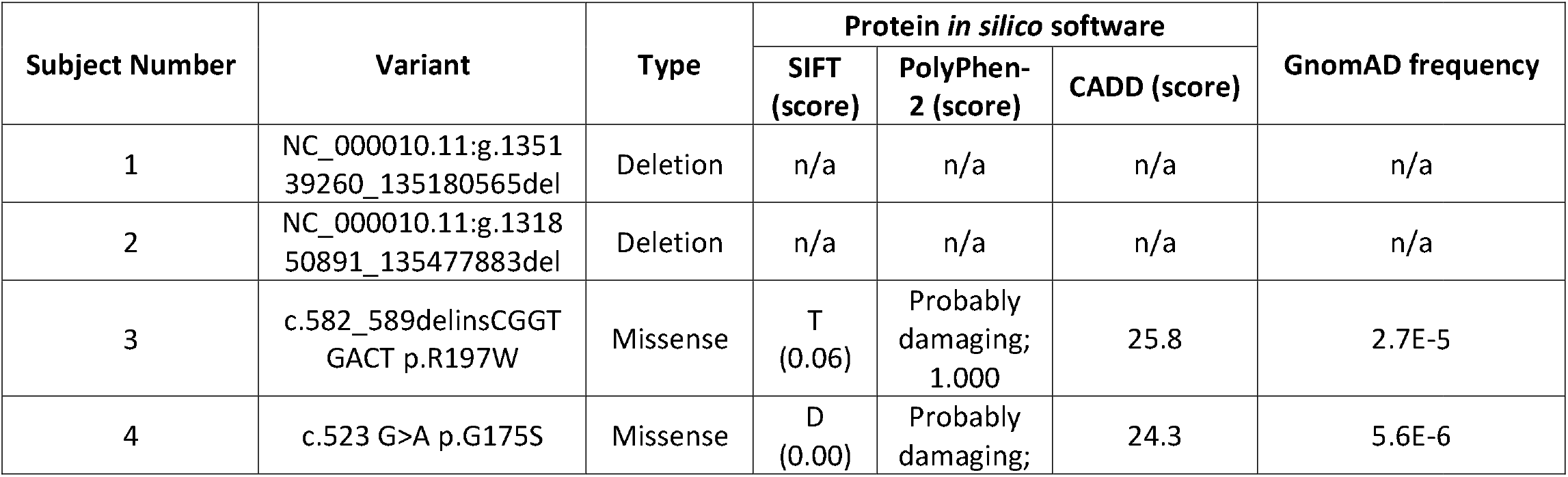
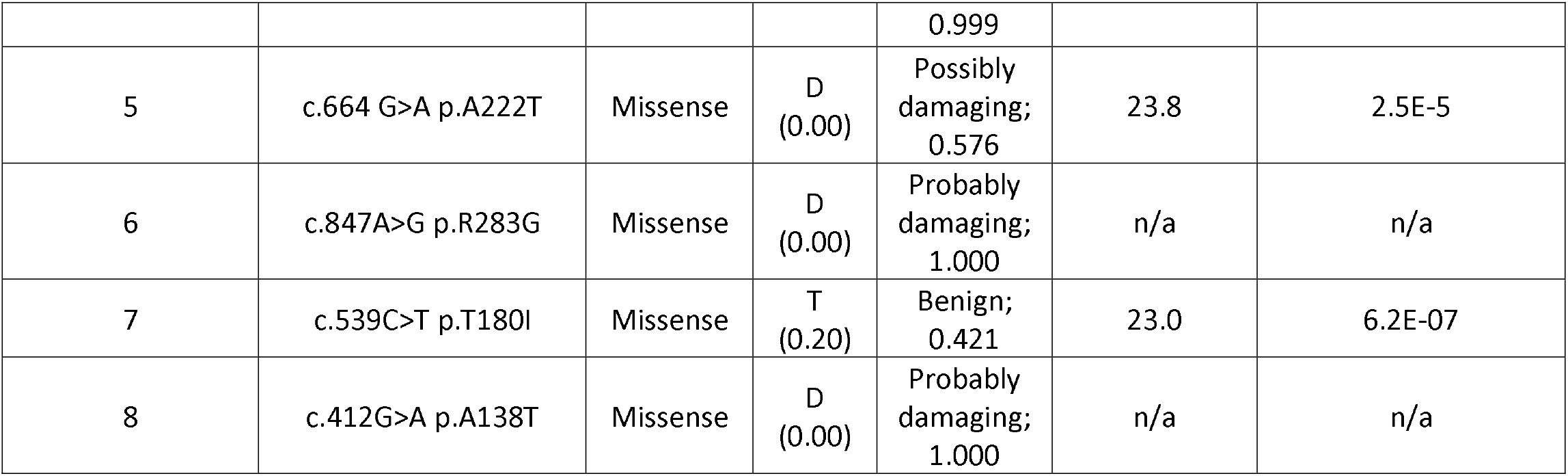
Pathogenicity data for novel *ECHS1* variants reported in our series.

Two previously published recurrent variants were also seen in this new cohort: the c.518C>T (p.A173V) variant and the c.489G>A (p.P163=, shown to reduce normal spliced RNA abundance) Pacific Islander/East Asian founder variant.^15, 23^

The c.518C>T (p.A173V) variant has been identified as being associated with paroxysmal exercise-induced dystonia.^15^ At the time of this review, 7 patients have been published in the literature with this variant and all 7 exhibit a form of paroxysmal exercise-induced dystonia.^16, 24, 25, 7, 15^ In our cohort of 26 newly reported patients, 5 have the c.518C>T (p.A173V) variant; all 5 have been reported to exhibit a form of paroxysmal exercise-induced dystonia. Out of these 5 patients, 1 patient is categorized in the infantile onset group and 4 patients are categorized in the late onset group.

To further explore genotype-phenotype correlation, we employed linear regression to investigate the effect of variant type on age of death or age at presentation and found that mutation severity or type is not associated with age of death or age at first symptoms. Furthermore, our analyses found no association between mutation category and binary/nominal clinical outcomes or diagnoses.

**Figure C.**
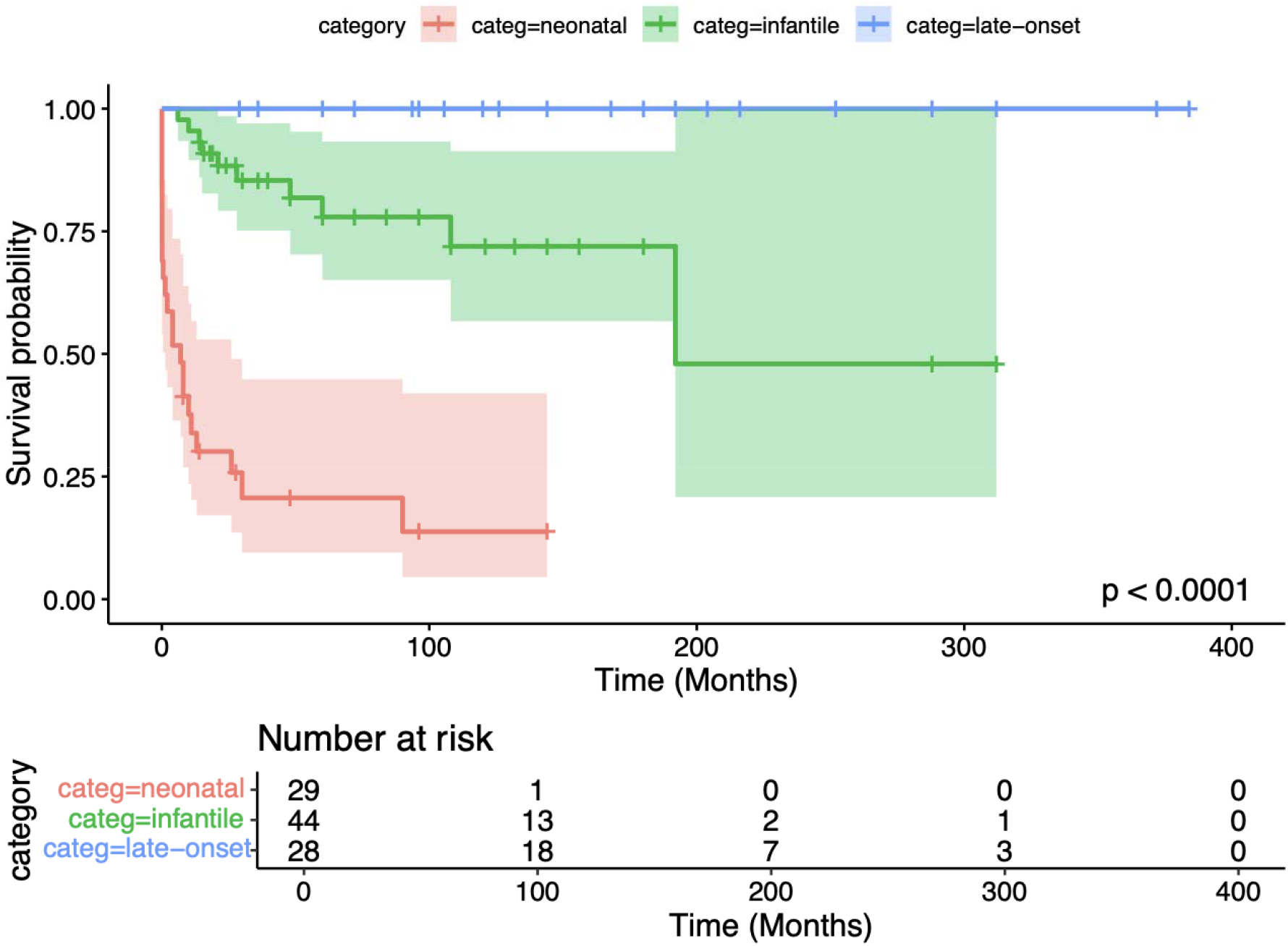
Kaplan-Meier curve demonstrating survival probability across three clinical groups. Table 2. Pathogenicity data for novel ECHS1 variants reported in our series

#### Clinical Presentation *Neonatal Presentation*

For the purposes of this study, subjects in the neonatal presentation group present within 30 days of birth.

We report 30 patients in this group (8 newly reported patients, 3 newly reported variants). ^16, 26, 14, 27, 28, 29^ 30/30 patients reported age of presentation: 27/30 patients presented immediately within 2 days of birth, 1/30 presented within 5 days of birth, and 2/30 presented within 30 days of birth. Of the 30 patients, 23 are deceased with a mean age of death of 9.36 months and a median age of death of 2.03 months.

The most common reported findings in this group were hypotonia (21/30), cardiomyopathy (18/30), lactic acidosis (17/30), developmental delay (14/30), presence of 2-methyl-2,3-dihydroxybutyric acid in UOA (11/21), seizures (11/30), deafness (11/30), and elevated lactic acid in UOA (10/21). Other reported findings included encephalopathy (7/30), feeding tube (7/30), dystonia (7/30), microcephaly (5/30), and ophthalmologic abnormality (5/30). Other potential biochemical markers include elevated alanine levels (9/21), 3-methylglutaconic acid (3/21), 2-hydroxybutyric acid (2/21), 3-hydroxybutyric acid (1/21), and S- (2-carboxypropyl)cysteine (1/1). Of note, 21 out ot 30 patients were recorded as having biochemical metabolite analysis performed in the neonatal onset group and only one patient was confirmed to have measured S-(2-carboxypropyl)cysteine.

#### Infantile Presentation

For the purposes of this study, subjects in the infantile presentation group present in between 30 days and 1 year after birth.

We report 45 patients in this group (8 newly reported patients, 3 newly reported variants). ^18, 30, 4, 17, 22, 31, 32, 20, 7, 33, 2, 21, 34, 23, 35, 12, 36, 37, 13^ 45/45 patients reported age of presentation: from these 45 patients there is a mean age of presentation of 6.7 months (median = 6 months). Of these 45 patients, 11 are deceased with a mean average age of death of 48.36 months and a median age of death of 28 months.

The most common reported findings in this group were developmental delay (36/45), dystonia (27/45), ophthalmologic abnormality (24/45), hypotonia (23/45), seizures (21/45), feeding tube (21/45), lactic acidosis (19/45), deafness (12/45), and elevated lactic acid level in UOA (13/29). Presence of 2-methyl-2,3-dihydroxybutyric acid exists (23/29) but other biochemical markers are also slightly more prevalent than in neonatal presentation – specifically, 3-methylglutaconic acid (10/29), elevated alanine levels (9/29), and 3-hydroxyisovaleric acid (8/29). Of note, 29 out ot 45 patients were recorded as having biochemical metabolite analysis performed in the infantile onset group. Cardiac findings were markedly decreased in this group – only one patient was reported to have cardiomyopathy compared to 18/30 in the neonatal presentation group.

Brain imaging and MRI results demonstrated varying findings such as caudate abnormality (26/45), globus pallidus abnormality (26/45), putamen abnormality (25/45), basal ganglia abnormality (12/45), and corpus callosum abnormality (4/45).

#### Late Onset

For the purposes of this study, subjects in the late onset group present after 1 year of age.

We report 38 patients in this group (10 newly reported patients, 3 newly reported variants). ^38, 30, 29, 22, 31, 24, 25, 7, 15, 6, 19, 5, 35, 12, 40, 41^ 33/38 patients reported age of presentation: from these 33 patients there is a mean age of presentation of 41.05 months (median = 26 months).

The most common reported findings in this group were dystonia (31/38), developmental delay (23/38), lactic acidosis (13/38), ophthalmologic abnormality (13/38), hypotonia (12/38), presence of 2-methyl-2,3-dihydroxybutyric acid in UOA (10/19), sensorineural deafness (11/38), and feeding tube (9/38).

More biochemical markers can be identified, including an elevated lactic acid level in UOA (7/19), 3-methylglutaconic acid (5/19), S-(2-carboxypropyl)cysteine (4/4), 3-hydroxyisovaleric acid (2/19), and 3-hydroxybutyric acid (2/19). Of note, 19 out ot 38 patients were recorded as having biochemical metabolite analysis performed and four patients were confirmed to have measured S-(2-carboxypropyl)cysteine in the late onset group.

Brain imaging and MRI results are significant for globus pallidus abnormality (27/38), putamen abnormality (15/38), caudate abnormality (13/38), and unspecified basal ganglia abnormality (4/38). One patient reported corpus callosum abnormality.

Treatment with n-acetylcysteine (7/38, 6/10 newly reported patients) or valine-restricted diet (5/38, 3/10 newly reported patients) was common in the newly reported patients.

### Neuroimaging

Neuroimaging data for 93 patients, 83 of whom were identified through a literature review, were centrally compared and reviewed by a pediatric neuroradiologist. Original imaging studies or MRI reports were available for 10 newly reported patients. Among these 10 patients, three presented in the neonatal period with two exhibiting abnormal morphology and a thin corpus callosum accompanied by diffusely reduced white matter volume. The other seven cases primarily showed features of Leigh syndrome, five of them with selective bilateral lesions involving the globus pallidus (Figure D).

**Figure D.**
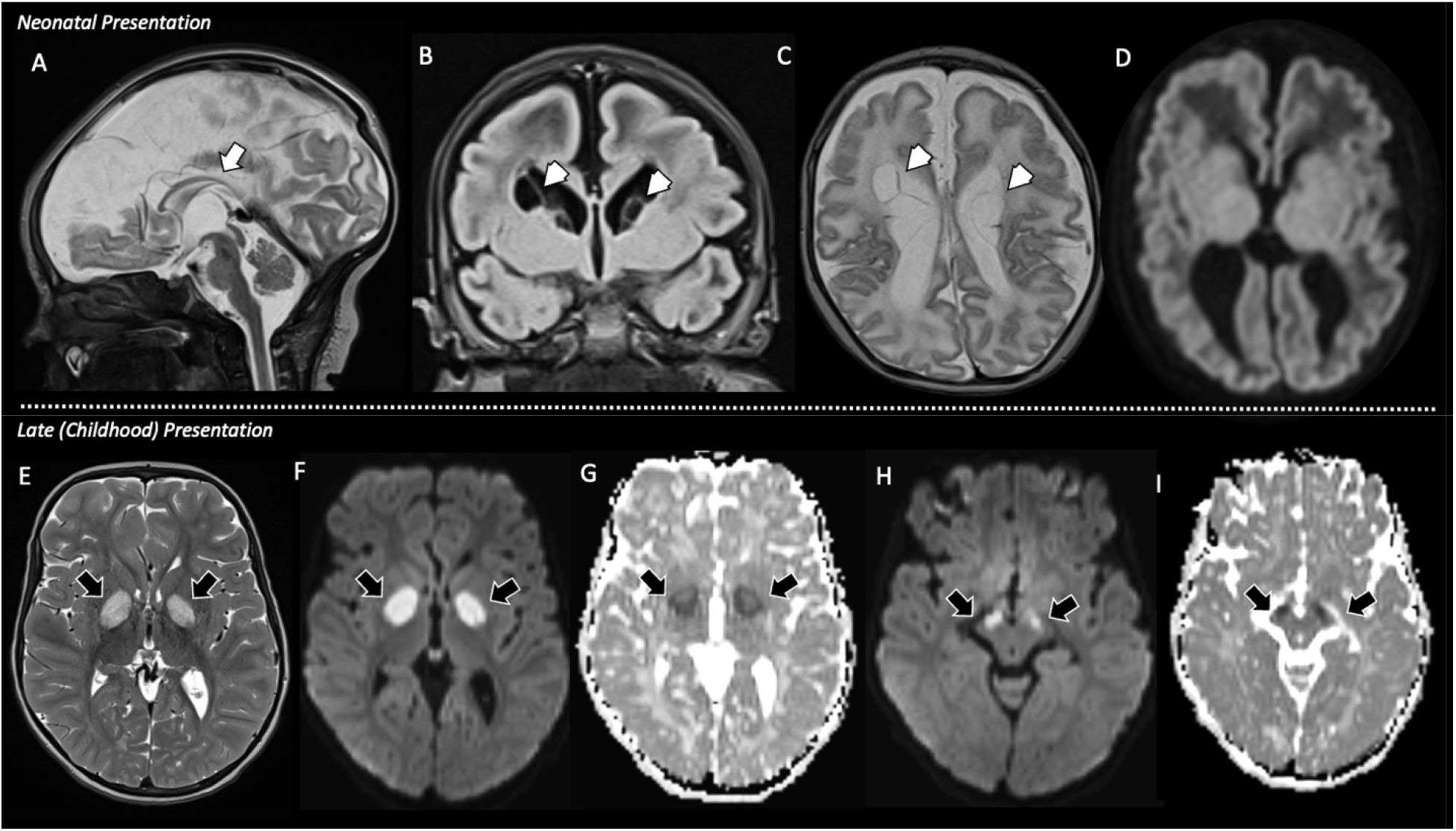
Brain MRI of two different patients: neonatal presentation (A–D) and late presentation (E–I). **Neonatal Presentation (A–D):** Midsagittal T2-weighted imaging shows severe hypoplasia of the corpus callosum (arrow, A). Coronal FLAIR and axial T2 images demonstrate multiple ventricular septations (arrowheads, B and C) without evidence of restricted diffusivity (D). **Late Presentation (E–I):** Selective lesions are observed bilaterally in the globus pallidus and substantia nigra, characterized by increased T2 signal (arrows, E). These findings are associated with reduced diffusivity, as confirmed on the Apparent Diffusion Coeffiicent (ADC) map (arrows, F–I).

**Figure E.**
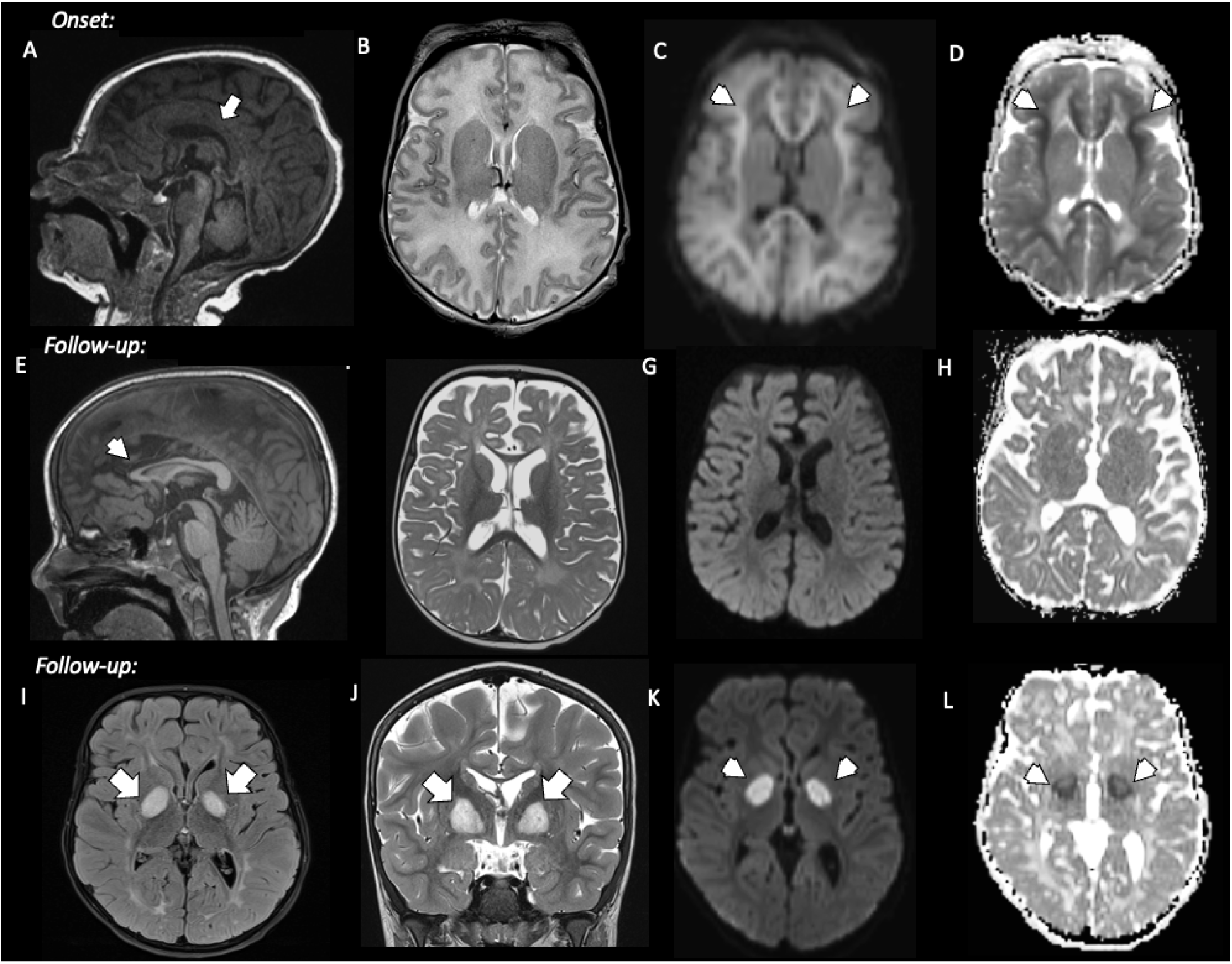
Longitudinal MRI study of a male patient with neonatal onset and late follow-up. **(A–D) Neonatal presentation:** (A) Sagittal T1-weighted image shows normal corpus callosum (arrow, A). (C, D) Diffusion-weighted and Apparent Diffusion Coefficient (ADC) map reveals extensive areas of reduced diffusivity involving the subcortical regions with relative preservation of the periventricular white matter; (arrowhead, C and D). **(E–H) First follow-up:** (E) Sagittal T1-weighted image shows atrophy of the corpus callosum, predominantly involving the genu and anterior body (arrowhead). (F) Axial T2-weighted image demonstrates mild residual white matter hyperintensity and associated volume loss. (G, H) Diffusion-weighted imaging and ADC maps show no areas of restricted diffusion. **(I–L) Late presentation:** (I) Axial FLAIR and (J) coronal T2-weighted images show new selective hyperintense lesions in the globus pallidus (arrowheads). DWI and corresponding ADC map confirm restricted diffusion within these lesions (arrowheads).

Follow-up MRI studies were performed for three patients. The first, a patient with neonatal onset, initially showed diffuse white matter changes with areas of reduced diffusivity in the superficial white matter as a newborn followed by a second suspected metabolic injury a few years later, where lesions selectively involved the globus pallidus bilaterally, along with residual changes noted in the white matter (Figure E). The second patient had a brain MRI study as a newborn, and a follow-up a few weeks later showed stable features, including white matter volume loss, intraventricular septations and enlarged lateral ventricles, and dysgenesis of the corpus callosum. The third patient presented as an infant with selective, symmetrical bilateral lesions in the globus pallidus, with subsequent imaging one year later demonstrating new lesions extending through the bilateral striatum, the mesencephalic tegmentum, and focal areas of the cerebellar hemispheres.

In the aggregate analysis of the 93 patients, structural abnormalities, including intraventricular septations, white matter volume loss, and dysgenesis of the corpus callosum, were present in 6.5% (n=6) of cases. Most of the cohort, 84.9% (79 of 93), presented with imaging features typical of Leigh syndrome, characterized by basal ganglia and/or brainstem involvement. Detailed anatomical distribution analysis was possible for 73 patients. Within this group, the entire basal ganglia (caudate, putamen, and globus pallidus) were diffusely affected in 45.2% (33 of 73) of patients while 37% (27 of 73) showed selective globus pallidus involvement. Furthermore, six patients (6.5%) who presented in the neonatal period exhibited diffuse white matter signal changes with an edematous appearance and components of reduced diffusivity, while only one patient was described as having a normal brain MRI study.

### Therapies

21.2% (N=24) of all patients were reported to be on diet therapy. Specifically, a valine-restricted diet was reported in 16 patients, the ketogenic diet was reported in 5 patients, and a low-protein diet was reported in 3 patients. Further dietary details were not provided. 19.5% (N=22) of all patients were on N-acetylcysteine; however, N-acetylcysteine was discontinued in 3 patients due to reported ineffectiveness or no improvement.^18, 23^ One patient in our newly-reported cohort with neonatal primary lactic acidosis was given sodium dichloroacetate via eIND, with favorable response. One patient in our newly-reported cohort is also on triheptanoin, a medium odd-chain triglyceride that has been used to treat other fatty acid oxidation disorders, and ECHS1D in one case report.^42^ While the previously published case reports moderate clinical improvement in their patient, the impacts in our reported patient are currently unclear.

### Survival, Functional Outcomes, and Mortality

As described above, statistical analysis revealed no association between genotype and survival. When we investigated survival with association to presentation category, as demonstrated in Figure C, we found that neonatal onset disease was associated with significantly increased risk of mortality when compared with later onset disease.

Among surviving patients, age at last follow-up differed by presentation category (Table 1). Patients with late onset disease were followed to a significantly greater age than those with neonatal or infantile presentation (pooled newly and previously reported cohorts: mean 152.79 months [SD 96.50; median 123], N=28, vs. 85.87 months [SD 67.29; median 72], N=42; Welch’s t-test, p=0.0026). There was no significant difference in age at last follow-up between the neonatal onset (mean 69.37 months, N=7) and infantile onset (mean 89.17 months, N=35) groups (p=0.3616). In contrast, late onset patients were significantly older at last follow-up than both neonatal onset (p=0.0031) and infantile onset (p=0.0054) patients. Interestingly, mean and median age at last follow-up were lower in the newly reported cohort than in the previously reported cohort for all three presentation groups (neonatal: mean 52.67 vs. 81.90 months, median 48 vs. 78 months; infantile: mean 60.86 vs. 96.24 months, median 36 vs. 84. months; late onset: mean 135.60 vs. 162.34 months, median 108 vs. 135 months), which may reflect earlier recognition and diagnosis of ECHS1 deficiency in more recent years.

Among neonatal onset patients with reportable data (n=5 out of 7), 5/5 were nonambulatory but developmentally appropriate for age, 3/5 required tube feeding, and 3/5 required invasive ventilatory support. However, these data from the neonatal onset group are not representative of the full clinical picture as early neonatal decompensation likely includes a conglomeration of life-saving measures.

In the infantile onset group (N=7 out of 8), outcomes were more heterogeneous: 5/7 were non-ambulatory and 4/7 tube-dependent, while 2/7 achieved independent ambulation with oral feeding and normal development.

Late onset patients (N=10 out of 11) had the most favorable functional trajectories overall. 7/10 achieved independent or assisted ambulation, 8/10 required no respiratory support, and 5/10 were fully orally fed. 3/10 remained non-ambulatory and tube-fed, underscoring that even “milder” late onset cases carry meaningful risk of significant disability.

Communication impairment (non-verbal status or loss of previously acquired expressive language) was documented in 6/22 patients, occurring after developmental regression in the infantile and late onset groups.

## DISCUSSION

To our knowledge, this is the largest known collection of individuals with ECHS1D described to date, comprising 87 patients from published reports and 26 newly reported patients across 12 institutions globally. Through new data identified in this case series, these data substantially add to the literature by broadening the phenotypic spectrum, refining natural history speculations, expanding on the clinical utility of biochemical markers, and identifying emerging improvement in contemporary survival trends, possibly associated with new potential therapies as described in our cohort.

Our findings demonstrate ECHS1D is a pan-ethnic disease, observed across all ethnicities, and without sex-specific differences in clinical characteristics. We report here the first known African American patient with ECHS1D. Notably, the African American child reported was misdiagnosed with cerebral palsy for the first several years of her life, highlighting the importance of maintaining a high index of suspicion in children with apparent cerebral palsy regardless of ancestry.

In keeping with established literature, our findings show that consistent genotype-phenotype correlations are not characteristic of ECHS1D, although some trends emerge. We present 8 newly described variants (31% of the newly reported patients in this cohort) and several recurrent variants have been identified. We corroborate that the c.518C>T p.A173V variant appears to be uniformly associated with paroxysmal exercise-induced dystonia, consistent with previously published reports of this variant.^16, 24, 25, 7, 15^ Additional features in this group included sensorineural hearing loss in 3 and developmental delay in one. While this group was biochemically more mild, with normal lactic acid in most cases, radiographic involvement of the basal ganglia, primarily the globus pallidus was still universal.

The c.489G>A p.P163= variant, reported in two of our patients as well as in the broader literature, appears to be associated with late onset disease. It has not yet been observed in neonatal or severe infantile phenotypes, possibly establishing itself as a hypomorphic allele. This variant was first reported as a founder variant in the Pacific Islands and now has been reported in additional populations in the East Asian population.^23^ The variant is thought to have high residual enzyme activity and does not cause disease in homozygotes but appears to cause a mild phenotype in patients when in the compound heterozygous state with a different pathogenic variant. One of these patients also had sensorineural hearing loss and one had developmental delay. All of the patients in our cohort with either the c.518C>T variant or the c.489G>A variant were categorized in the late onset group. These data strongly suggest that the third class of ECHS1D, which we have called “late-onset”, but has alternatively been called “paroxysmal dystonia subtype” is distinct in three ways: genotypically, phenotypically and by age of onset, while being radiographically highly similar to the other groups. Across this subgroup, dystonia was universal, and sensorineural hearing loss was common. There exist in this subgroup additional patients with neither of the recurrent genotype, although all such patients had developmental delay.

We speculate that this subgroup has high residual enzyme activity, but the exact genotypic or enzymatic definition of this group is still not known, nor is the full spectrum of disease.

In our cohort, no statistically significant survival difference between missense vs. predicted LOF variants could be identified, suggesting that clinical course may also depend on secondary modifier effects, functional enzyme activity, or other factors beyond gene disruption alone. Unfortunately, enzyme activity measurements were not available on almost any patients. Furthermore, we found no association between a particular genotype and the presence or absence of clinical characteristics or specific organ involvement. These findings should not be taken to mean that genotype is uninformative in ECHS1D. While age at onset provides a broadly applicable first-tier estimate of disease severity and survival, genotype may be able to refine that estimate for the subset of patients carrying alleles of established functional consequence (most notably the paroxysmal dystonia phenotype). A limitation of our variant analysis is that pathogenicity of the eight newly reported variants was assigned using ACMG criteria supported by *in silico* prediction and population frequency data without accompanying enzymatic evidence. While ideally newly identified *ECHS1* variants should ideally undergo functional validation using strategies such as those described by Kishita et al.,^43^ it is important to note that variants of uncertain significance are common in *ECHS1* and that functional testing is limited in availability.

Biochemical findings did not appear in patients consistently, limiting their clinical utility as diagnostic or prognostic biomarkers. Biochemical markers trended toward greater specificity than sensitivity, with greater utility when positive. Of these, 2-methyl-2,3-dihydroxybutyric acid was present approximately 50-80% of the time across all groups (11/21 neonatal, 23/29 infantile, 10/19 late onset). Other biochemical findings included 3-methylglutaconic acid, which appeared most commonly in the infantile cohort (reported as positive 10/29 times, or 34.5% of the time), hyperlactatemia which appears in all phenotypic cohorts but is most common in neonatal presentations (10/21 times, 47.6%), and alanine elevation, which is likely a reflection of impaired lactate metabolism. Other minor markers identified include 3-hydroxyisovaleric acid (observed mostly in infantile onset), S-(2-carboxypropyl) cysteine (observed mostly in late onset; however, may have been underascertained), and 2-hydroxybutyric acid or 3-hydroxybutyric acid. These data suggest a supportive role of a small molecule work up in patients with suspected or proven ECHS1 deficiency, for clinical care or work up of variants of unknown significance. These data also continue to support the underlying hypothesis that while ECHS1 participates in both mitochondrial fatty acid β-oxidation as well as valine metabolism, impaired valine metabolism is a major driver in pathogenesis. ECHS1 metabolizes methacrylyl-CoA, a reactive intermediate of valine catabolism; when ECHS1 activity is reduced, methacrylyl-CoA and related intermediates accumulate and can react with mitochondrial thiol-containing proteins, including the lipoyl domain of the private dehydrogenase complex, thereby impairing energy metabolism. Although the absence of acylcarnitine abnormalities and a typical fatty acid oxidation phenotype suggests that impaired β-oxidation is not the primary disease driver, ECHS1 also functions in short-chain fatty acid β-oxidation, and disruption of this pathway may contribute to secondary OXPHOS dysfunction and disease severity.^44^

Our cohort affirms that ECHS1D presents along a broad clinical spectrum, which we have categorized into neonatal, infantile, and late-onset groups based on age of onset and clinical course. These groupings demonstrated distinct trends in presentation, imaging findings, survival, and treatment response.

Neonatal-onset cases were characterized by early and severe multisystem disease, often with lactic acidosis, hypotonia, cardiomyopathy, and high early mortality (62.5%). Although we did not collect the data in a structured way, five of the patients in the neonatal cohort were known to have pulmonary hypertension, which may have been secondary to lactic acidosis and contributes significant to morbidity and mortality. In previous literature, cardiomyopathy has been reported as dilated or hypertrophic and occurring in about 60% of patients, with transient cardiac hypertrophy reported in two cases.^4, 17^ We found that variations of structural cardiac congenital abnormalities and cardiomyopathy are widely reported in the neonatal presentation group. Median age of death in this group was 1.66 months, highlighting the aggressive nature of this phenotype. In contrast, late-onset patients, who typically presented after one year of age, had more localized neurologic symptoms, particularly dystonia, and significantly lower mortality (0%). Surviving late-onset patients were also followed to a significantly greater age than neonatal and infantile onset patients, which together with their lower mortality, suggests a milder disease course and perhaps better effectiveness of therapeutic intervention.

The infantile-onset group was heterogeneous, but many exhibited hallmark features such as dystonia, developmental delay, and basal ganglia involvement on neuroimaging, in line with existing data signaling ECHS1D as a common etiology of Leigh syndrome. Although mortality was lower than in the neonatal group, a subset of patients did experience disease progression and early death. Later disease onset is also correlated with more favorable functional outcomes related to ambulation, respiration, feeding, and overall disability.

Similar to other inborn errors of amino acid metabolism, there are individual reports of very acute decompensations in previously healthy children in the setting of infection.^45^ This suggests that the approach to managing ECHS1D may ultimately borrow from its related amino acid organic acidopathy conditions, with an eventual goal of structured therapeutic and dietary recommendations and newborn screening approaches. Of note, the reported ability of some patients with ECHS1D to tolerate ketogenic diet highlights that it is likely the impaired amino acid catabolism and not the impaired fatty acid oxidation defect that contributes to disease, although the data remain limited. These findings highlight that ECHS1D represents a spectrum disease, with limited genotype-phenotype correlation, contributing to variability in outcomes. Nonetheless, families and providers often seek prognostic information in the face of a challenging and rare diagnosis. Therefore, categorization by age of onset does seem to be a meaningful way to provide insight on predicted morbidity and mortality.

The neuroimaging findings in our cohort and literature review highlight a significant phenotypic overlap with the radiological features associated with pathogenic variants of the Pyruvate Dehydrogenase Complex (PDHC), including structural malformations or Leigh syndrome features.^46, 47^ Structural abnormalities often identified in the prenatal period of patients with PDHC,^48^ specifically the dysgenesis of the corpus callosum, intraventricular septations, and significant white matter volume loss, were noted in 6.5% of the ECHS1D patients. These neuroimaging features are hallmark indicators of potential early metabolic failure that disrupts normal brain development in utero in some ECHS1D patients.

During childhood, most patients presented with Leigh syndrome imaging features, characterized by predilection for the basal ganglia lesions, particularly selective involvement of the globus pallidus. In our aggregate analysis, 84.9% of patients exhibited features typical of Leigh syndrome, with selective globus pallidus involvement appearing in 37% of those cases. One newly described patient with initial imaging revealing changes selective of the globus pallidus later developed into diffuse basal ganglia findings on follow-up imaging.

Interestingly, a small subset of our cohort and literature review (6.5%) presented with acute neonatal injury characterized by diffuse subcortical white matter signal changes and components of reduced diffusivity. These features are not specific, but we believe it may reflect an early disease state prone to superimposed systemic complications in the context of, for example, primary lactic acidosis often noted at this age. One notable neonatal patient in our series initially presented with these diffuse white matter changes and subsequently developed new disease activity later in childhood with imaging features characteristic of Leigh syndrome, including selective globus pallidus involvement in association with the residual white matter changes from prior injury. This progression suggests that the presence of superimposed mild white matter signal changes in ECHS1D*-*related patients, currently presenting with Leigh syndrome features, may be at least partially explained by a prior, and potentially subclinical for some cases, disease onset during the neonatal period. These findings underscore the importance of recognizing the transition from early, diffuse white matter injury to localized, metabolic driven deep gray matter lesions as the disease course unfolds.

Overall survival was 77% in our newly reported cohort (N = 26) compared with 69% in previously reported patients (N = 87). Moreover, we report a 37% survival rate in newly reported neonatal onset patients compared with 18.2% in prior reports, possibly reflective of earlier recognition of ECHS1D, improvements in NICU care, and we speculate due to greater utilization of therapies. Several therapies were reported in our cohort, including valine-restriction most commonly, N-acetylcysteine, and investigational eIND use of sodium dichloroacetate which appeared to stabilize metabolic acidosis in one case of neonatal ECHS1D with primary lactic acidosis. perform a formal treatment-versus-outcome comparison over our retrospective review and at this time, the data are insufficient to propose one form of therapy over another. Further prospective clinical study regarding the utility of specific therapies is needed to ascertain treatment efficacy. An individualized approach, or a combination of various treatment modalities, may be proposed for empiric treatment of ECHS1D.

Our study is limited by being a retrospective, multi-center study with heterogeneity in the data provided across centers. Although the retrospective, multi-center nature has enabled capture of a much larger number of patients than have previously been co-analyzed, it is intrinsically limited in its ability to provide nuanced functional data and treatment responses. Because treatments were not randomized or placebo-controlled, and treated patients differed from untreated patients in severity with multiple potential confounders, this study cannot support causal inference regarding any therapy; all treatment-related observations are hypothesis-generating. Similarly, the heterogeneity in neuroimaging limits our ability to draw conclusions. Nonetheless, this work serves as a foundation for designing future prospective natural history & therapeutic studies.

## Data Availability

All data produced in the present study are available upon reasonable request to the authors.

## ACKNOWLEDGMENTS

Special thanks to Dora Lacarrubba.

## TAKE-HOME

In the largest reported cohort of ECHS1 deficiency to date (113 patients), age of onset—neonatal, infantile, or late-onset—predicts disease severity and mortality more reliably than genotype, with two recurrent variants associated with a distinct, milder, paroxysmal dystonia phenotype.

## AUTHOR CONTRIBUTIONS

Conceptualization: I.E., R.G., C.K. Data curation: C.A., I.E., R.G., C.K.

Formal analysis: C.A., I.E., R.G., C.K., R.X.

Funding acquisition: n/a Investigation: I.E., R.G., C.K.

Methodology: I.E., R.G., C.K. Project administration: n/a Resources: n/a

Software: R.G., R.X. Supervision: I.E., R.G. Validation: n/a

Visualization: C.A., R.G., C.K., G.V.

Writing – original draft: C.A., I.E., R.G., C.K.

Writing – review & editing: I.A., G.B., J.F.C., V.C., A.C.F., G.M.E., M.G., C.H., S.H., M.I., E.J., A.K., J.K., D.L., A.L., C.L., M.A., R.L., M.L., J.M., M.M., E.N., L.P., E.P., M.S., M.C.S., R.P.S., E.S., C.G.T., J.L.K.V., G.V., R.X.

## ETHICS APPROVAL/CONSENT

This study was evaluated by the Children’s Hospital of Philadelphia Institutional Review Board (IRB #23-021081) and determined to meet the criteria for exemption under 45 CFR 46.104(d)(4)(ii). A waiver of HIPAA authorization was granted per 45 CFR 164.512(i)(2)(ii) for access to identifiable information from medical records. All patient data were recorded in a deidentified manner. For unpublished cases contributed by collaborating institutions, ethical approval and data-sharing agreements were obtained from each participating center in accordance with local institutional guidelines. Informed consent was waived in accordance with the IRB exemption determination.

## AVAILABILITY OF DATA AND MATERIAL STATEMENT

The data that supports the findings of this study are available in the supporting information of this article.

## GENERATIVE AI

Generative AI was not used in any capacity for this manuscript.

